# Edge-tuning of artificial intelligence improves diagnostic performance for *Schistosomiasis haematobium* in a rural setting of Côte d’Ivoire

**DOI:** 10.1101/2025.07.11.25331398

**Authors:** María Díaz de León Derby, Jean T. Coulibaly, Elena Dacal, Kigbafori D. Silue, Daniel Cuadrado, David Bermejo-Peláez, Jaime García-Villena, Lin Lin, Karla N. Fisher, Jason R. Andrews, Daniel A. Fletcher, Miguel Luengo-Oroz, Isaac I. Bogoch

**Affiliations:** Department of Bioengineering, University of California, Berkeley, Berkeley, California; Unité de Formation et de Recherche Biosciences, Université Félix Houphouët-Boigny, Abidjan, Côte d’Ivoire; Centre Suisse de Recherches Scientifiques en Côte d’Ivoire, Abidjan, Côte d’Ivoire; Spotlab, Madrid, Spain; Biomedical Image Technologies, ETSI Telecomunicación, Universidad Politécnica de Madrid, Madrid, Spain; CIBER de Bioingeniería, Biomateriales y Nanomedicina, Instituto de Salud Carlos III, Madrid, Spain; Biological Systems and Engineering Division, Lawrence Berkeley National Laboratory, University of California, Berkeley, Berkeley, California; Chan Zuckerberg Biohub, San Francisco, California; Divisions of General Internal Medicine and Infectious Diseases, Toronto General Hospital, University Health Network, Toronto, Canada; Department of Medicine, Stanford University School of Medicine, Stanford, CA; Department of Medicine, University of Toronto, Toronto, Canada

**Author notes:** Contributed equally to this manuscript (María Díaz de León Derby, Jean T. Coulibaly, Elena Dacal). **Corresponding Authors:** Isaac I. Bogoch, Divisions of General Internal Medicine and Infectious Diseases, Toronto General Hospital, 14EN 209, 200 Elizabeth Street, Toronto, ON, Canada M5G 2C4.; Miguel Luengo Oroz, Spotlab, Madrid, Paseo Juan XXIII, 36B, 28040, Spain.; Daniel Fletcher, Stanley Hall, University of California, Berkeley. Berkeley, CA, 94720. United States.

## Abstract

**Background:** Schistosomiasis affects over 200 million people and causes significant urogenital and gastrointestinal morbidity. Mass drug administration (MDA) with praziquantel is used to mitigate severe illness and reduce infection rates. Portable microscopy, combined with artificial intelligence (AI), offers a novel method for schistosomiasis screening in low-resource settings. This study tested whether re-training AI models for *Schistosoma* egg detection with local field data, a process we call “edge-tuning”, could improve the model’s performance on the following field day.

**Methods:** This study in Côte d’Ivoire evaluated a portable microscope (NTDscope) for *Schistosoma haematobium* screening. Urine samples from 100 community members were analyzed using AI models on the NTDscope and traditional light microscopy. Starting AI models, trained on images from a previous version of the NTDscope, were edge-tuned after the first day of sample collection using cloud-based image annotation and re-training. Starting and edge-tuned models were evaluated at confidence thresholds optimizing for sensitivity, specificity, or egg counting.

**Findings:** For all thresholds, edge-tuned models performed better than starting AI models. Compared to manual counting of eggs on the NTDscope, sensitivity of the starting AI model on day 2 ranged from 59.3%-75.5%, with specificity ranging from 46.7%-85.7%. After edge-tuning, sensitivity increased to 77.8%-100%, with specificity from 78.6%-100%. Compared to light microscopy, edge-tuned AI models had comparable performance to manual counting from NTDscope images.

**Interpretation:** Portable microscopy is an effective solution for rapid, on-site schistosomiasis screening. AI-based egg detection increases diagnostic throughput while maintaining good performance. This study demonstrates that edge-tuning AI models with local data significantly improves their performance and can be performed in low-resource settings, making the combined technologies effective tools for monitoring schistosomiasis programs in endemic areas.

## Introduction

Schistosomiasis continues to impact over 200 million people globally, disproportionately affecting those who live in impoverished rural settings, particularly African children. This helminth infection is acquired through direct contact with contaminated fresh water and over 90% of the global burden of infection is in African settings.^1^ The infection is associated with considerable urogenital (*S. haematobium*) and gastrointestinal (*S. mansoni*) morbidity.

Mass drug administration (MDA) programs administer praziquantel at the community level on an annual or semi-annual basis to decrease the burden of infection and lessen morbidity.^2^ Despite the implementation of these programs in several African countries, it is crucial to ensure MDA is achieving its intended objectives, and in 2021, the World Health Organization (WHO) launched diagnostic target product profiles for the “monitoring, evaluation and surveillance of schistosomiasis control programmes.^3^”

Portable microscopy, combined with artificial intelligence (AI) models, is emerging as an effective method to screen for schistosomiasis and other neglected tropical diseases (NTDs).^4–10^ These devices may aid in identifying communities where MDA should be conducted and in monitoring the effectiveness of existing control programs. Here, we evaluate the utility of a portable microscope, the NTDscope, coupled with AI models for the screening of *S. haematobium* infections. We test whether an AI model trained on previous datasets performs better after being re-trained in real-time using data collected in a new testing location through a technique we call *edge-tuning*. By comparing the performance of the starting AI model to the edge-tuned AI model on the second day of our field study, we show that edge-tuning results in improvements in patient-level diagnostic sensitivity and specificity, as well as in egg-level quantification. When compared to light microscopy, the best-performing edge-tuned AI model achieved performance equivalent to manual counting of eggs in patient images on the NTDscope.

## Methods

### Location and REB

This study was conducted around the town of Azaguié, Côte d’Ivoire in January of 2024. Ethical approval was granted by the University Health Network, Toronto, Canada (REB #21-5582) and the Comité National d’Éthique des Sciences de la Vie et de la Santé, Abidjan, Côte d’Ivoire (REB #186-21). Ethical permission was also granted by the local health district officer.

### Sample collection and light microscopy

Approximately 100 community members aged five and older were asked to provide a single urine sample on two consecutive days. Adults provided written consent, and assenting children who had written consent from a parent or guardian were included. All who were found to be positive for *S. haematobium* were treated with praziquantel (40 mg/kg), as per country guidelines, free of charge. Urine samples were collected between 10:00 and 14:00 and processed on the day of collection. Urine containers were first shaken, and then 20 mL of urine was extracted using a plastic syringe; of this, 10 mL was separated and evaluated by conventional light microscopy, and 10 mL was separated and evaluated by portable microscopy using the NTDscope^11^ (Figure 1a). For light microscopy, as per WHO guidelines, 10 mL of urine was pressed through filter paper with 20-micron pores (Sterlitech PCTE filters; Washington, USA)^12^. Filter paper was then placed on a standard glass microscope slide with the application of one drop of Lugol’s iodine. Microscopists evaluated the slide under 10x and 20x objective lenses, recorded the presence or absence of *S. haematobium* eggs, and quantified egg intensity if present. 10% of samples were re-evaluated for quality control.

**Figure 1:**
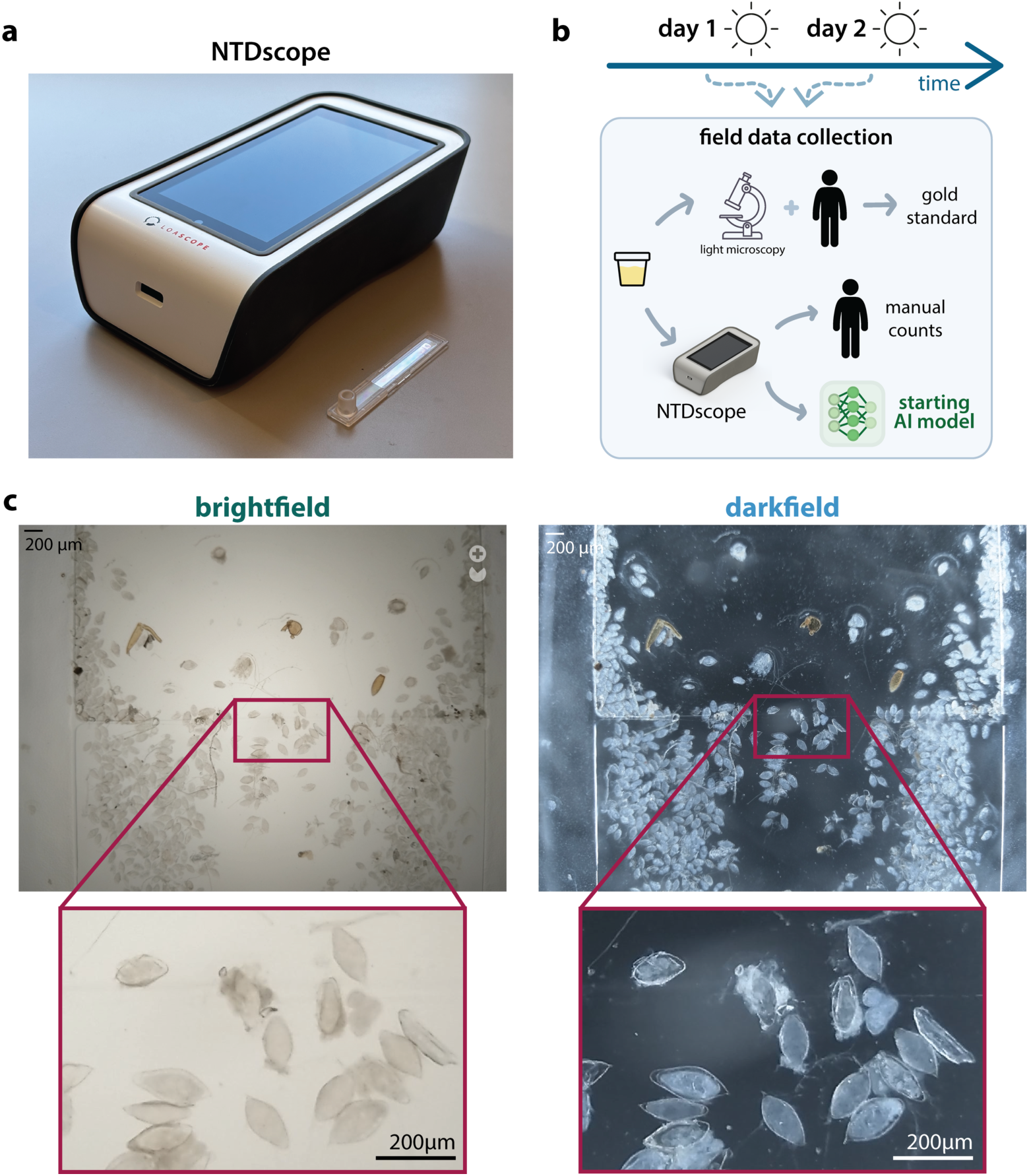
NTDscope and AI models can be used for schistosomiasis diagnosis in a rural setting in Côte d’Ivoire. **a.** Image of the NTDscope and a capillary used for urine sample processing. **b.** Field study timeline and data collection steps. **c.** Examples of a brightfield and darkfield image acquired during the field study, with an inset showing S. haematobium eggs.

### Capillary loading and NTDscope imaging

Based on initial results from light microscopy, we preferentially selected urine samples for re-evaluation using the NTDscope. For day 1, we selected 25 samples with high-intensity infection (greater than 50 eggs per 10 mL of urine), 25 samples with low-intensity infection (less than 50 eggs per 10 mL of urine), and 25 samples with 0 eggs.^12^ For day 2, we selected 24 samples with high-intensity infection (greater than 50 eggs per 10 mL of urine), 21 samples with low-intensity infection (less than 50 eggs per 10 mL of urine), and 23 samples with 0 eggs. Urine containers were once again shaken, and 10 mL of urine was extracted and pressed through an injection-molded plastic tapering capillary designed to trap *S. haematobium* eggs.^13,14^ The capillary was then wiped clean and inserted into the NTDscope for imaging (Figure 1b). Both brightfield and darkfield illumination were used to capture images in five fields-of-view (FOV). A representative FOV, imaged in brightfield and darkfield, is shown in Figure 1c.

### AI model training

The starting AI model is based on YOLOv8 pre-trained on the COCO 2019 dataset^15^. It was trained to perform an object detection task, which involved identifying *S. haematobium* eggs from images of patient urine samples, using images captured by a previous version of the NTDscope during two prior field studies in *S. haematobium*-endemic regions of Côte d’Ivoire.^13,14^

Two versions of the starting AI model were trained separately for images acquired in brightfield and darkfield illumination, respectively. A total of 526 (263 brightfield and 263 darkfield) images with 7811 labels (3659 brightfield and 4152 darkfield) were used for training.

Considering the trade-off between performance and latency, we selected the YOLOv8-small model with an input size of 1024 pixels. The models were trained using the AdamW optimizer, configured with a learning rate of 0.002, a first momentum parameter of 0.9, and a second momentum parameter of 0.999. Training was conducted with a batch size of 16 over 200 epochs, with early stopping implemented to stop training if performance did not improve over 20 consecutive epochs. Model training takes approximately one hour on a single GPU of NVIDIA A10G.

The AI model assigns a confidence score to each detection. This number, ranging from 0-1, indicates the model-assessed likelihood that the retrieved object is an egg. In practice, after a model is trained, a confidence score threshold is selected to decide whether the detections in new images are counted as eggs or not. For this study, three different confidence score thresholds were selected for each AI model, chosen to optimize one of the following: (i) patient-level sensitivity, (ii) patient-level specificity, and (iii) egg quantification accuracy. These thresholds were selected based on model performance on the validation set. For each sample acquired that was run through the trained models, the three thresholds were applied to quantify the number of AI detections per patient per image contrast (brightfield and darkfield). The results for all AI models (brightfield and darkfield versions of the starting and edge-tuned models) and thresholds are found in Tables 1 and 2.

**Table 1:** Performance of brightfield AI models compared to manual counts on NTDscope brightfield images.

| Day | AI model | Threshold | Patient-level |  | Egg-level |
| --- | --- | --- | --- | --- | --- |
|  |  |  | Sensitivity | Specificity | r |
| 1 | Starting model | Egg-quantification | 94.5 % | 65.0% | 0.872 |
| 1 | Starting model | Sensitivity-optimized | 98.2% | 15.0% | 0.879 |
| 1 | Starting model | Specificity-optimized | 70.9% | 80.0% | 0.847 |
| 2 | Starting model | Egg-quantification | 75.5% | 60% | 0.873 |
| 2 | Starting model | Sensitivity-optimized | 75.5% | 46.7% | 0.877 |
| 2 | Starting model | Specificity-optimized | 75.5% | 80.0% | 0.852 |
| 2 | Edge-tuned model | Egg-quantification | 100% | 80.0% | 0.943 |
| 2 | Edge-tuned model | Sensitivity-optimized | 98.1% | 100% | 0.920 |
| 2 | Edge-tuned model | Specificity-optimized | 81.1% | 100% | 0.861 |

**Table 2:** Performance of darkfield AI models compared to manual counts on NTDscope darkfield images.

| Day | AI model | Threshold | Patient-level |  | Egg-level |
| --- | --- | --- | --- | --- | --- |
|  |  |  | Sensitivity | Specificity | r |
| 1 | Starting model | Egg-quantification | 68.4% | 88.8% | 0.822 |
| 1 | Starting model | Sensitivity-optimized | 91.2% | 50.0% | 0.809 |
| 1 | Starting model | Specificity-optimized | 40.4% | 100% | 0.699 |
| 2 | Starting model | Egg-quantification | 70.4% | 78.6% | 0.558 |
| 2 | Starting model | Sensitivity-optimized | 72.2% | 64.3% | 0.680 |
| 2 | Starting model | Specificity-optimized | 59.3% | 85.7% | 0.397 |
| 2 | Edge-tuned model | Egg-quantification | 94.4% | 78.6% | 0.965 |
| 2 | Edge-tuned model | Sensitivity-optimized | 90.7% | 100% | 0.961 |
| 2 | Edge-tuned model | Specificity-optimized | 77.8% | 100% | 0.911 |

### In-device inference pipeline

To enable on-site inference and counting without requiring an internet connection, the models were executed directly on the NTDscope^16^. The trained models were converted to TensorFlow Lite and fed through TensorFlow’s Android backend. Each of five FOV images taken with the scan routine described above were converted into input tensors for the models using a custom pipeline written in Kotlin, with computationally intensive calculations such as colorspace translation and image resizing optimized in C. This process yielded an average camera-to-inference time of less than 3 seconds per image on the NTDscope, which is powered by a low-mid range Qualcomm SDM660-based chipset^11^.

### S. haematobium egg labelling and quantification

Egg quantification was performed both automatically, with the AI models integrated with the NTDscope software, and manually from images acquired on the NTDscope. Images were uploaded via the mobile network to a telemicroscopy platform, Telespot (Spotlab, Madrid), which allowed for remote annotation of the images. Study personnel quantifying images were blinded to both light microscopy and AI results.

### Edge-tuning

Re-training of the starting AI model and an update to confidence score thresholds for the edge-tuned AI model was performed between day 1 and day 2 of sample collection to test for more accurate *S. haematobium* egg detection on day 2. The patient sample images acquired on day 1 were uploaded to Telespot and made accessible to remote annotators, who annotated all *S. haematobium* eggs present in the images and used these to re-train the brightfield and darkfield versions of the starting AI model. Over a span of 4 hours, seven labelers worked in parallel with the Telespot platform and analyzed 36 patient samples, identifying 190 positive images (96 brightfield, 94 darkfield) and generating a total of 7,900 new annotations (3,838 brightfield, 4,062 darkfield) (Figure 2).

**Figure 2:**
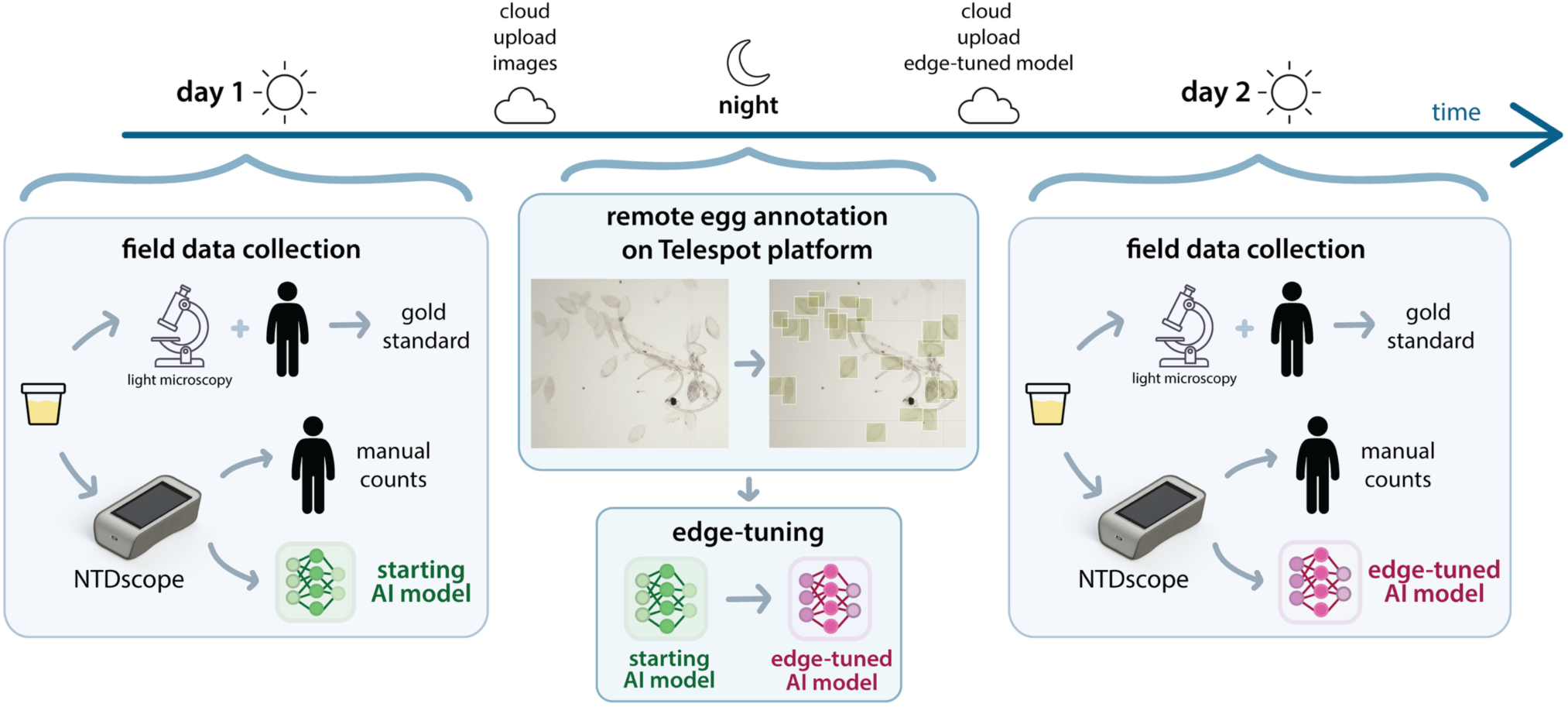
Timeline of 2-day field study in rural Côte d’Ivoire showing remote image annotation and edge-tuning, performed remotely at night between two field study days.

With this newly annotated dataset and the dataset used to train the previous models, the AI model was re-trained, requiring approximately two hours of training time. The images acquired on day 1 were split into training and validation sets, and therefore used to select new confidence score thresholds that maximized performance on the validation set. These thresholds were applied to the images analyzed by the edge-tuned AI model on day 2. The brightfield and darkfield versions of the edge-tuned AI models and updated confidence score thresholds were sent to the NTDscope via the Telespot platform, which is automatically synchronized with the app running on the NTDscope devices, and were used on all samples collected on day 2 (Figure 2). The samples collected on day 2 were also run through the starting AI model and compared with the edge-tuned model, with the aim of evaluating the usefulness of the edge-tuning technique (Figure 3).

**Figure 3:**
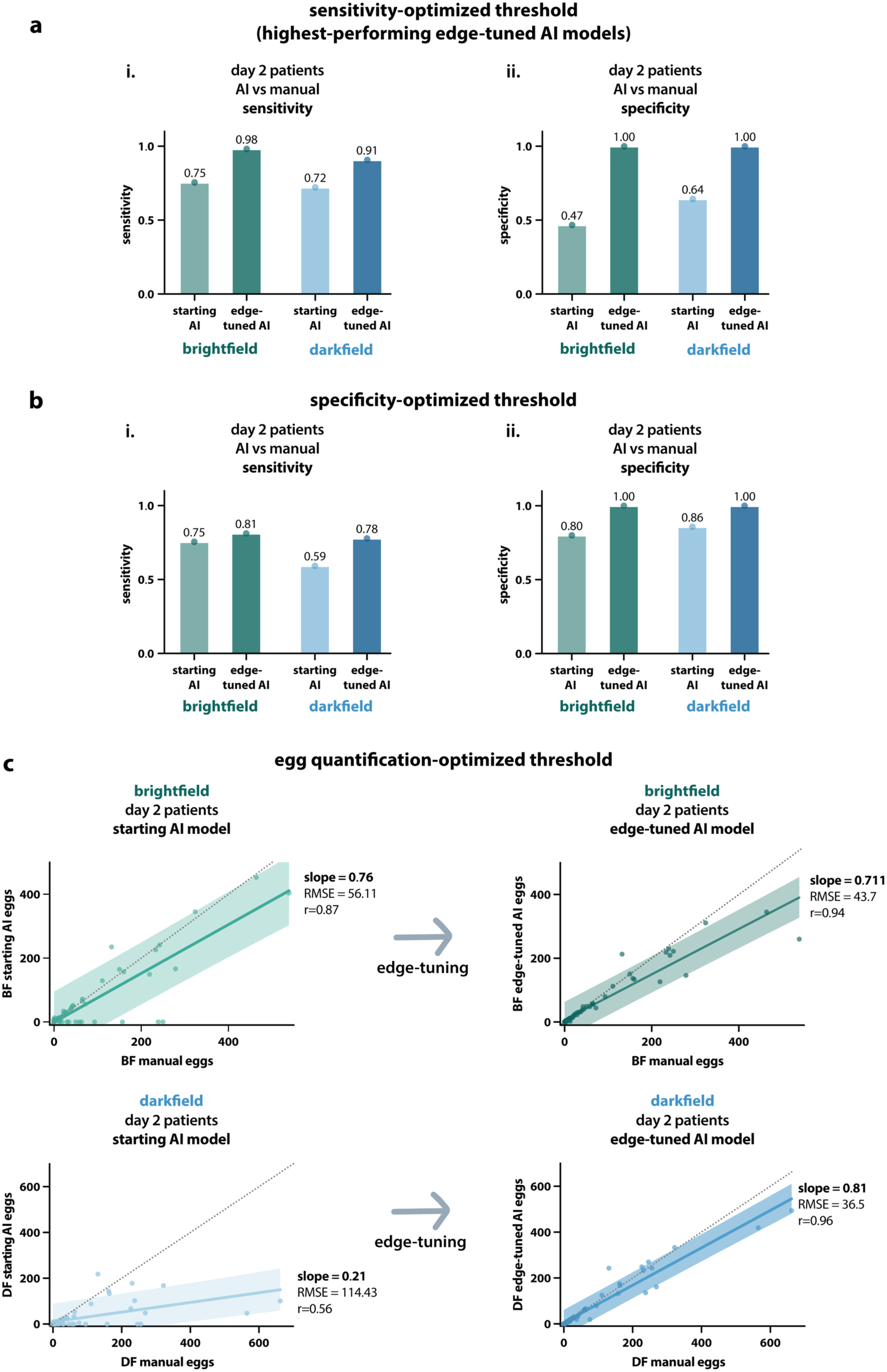
Edge-tuning improves performance on AI models and thresholds optimized for different metrics. All data in this figure is on patients from day 2 and shows results for starting AI models and edge-tuned AI models. **a.** Performance of AI models using the sensitivity-optimized confidence score threshold. Showing sensitivity (i) and specificity (ii) of the models using manual counts on NTDscope images as a reference. **b.** Performance of AI models using the specificity-optimized confidence score threshold. Showing sensitivity (i) and specificity (ii) of the models using manual counts on NTDscope images as a reference. **c.** Improvements in AI models with egg quantification-optimized confidence score thresholds, comparing patient-level egg counts by the AI models with manual egg counts on the NTDscope images. Plots show the slope of the linear fit to the data, the root mean square error (RMSE), and Pearson’s r for each model. Data is for brightfield (top) and darkfield (bottom) models.

### Statistical analysis

Results were entered into a Microsoft Excel file (Microsoft, Redmond, WA) and analyzed using R, version 4.1.3. We calculated sensitivity and specificity for egg detection, along with 95% exact binomial confidence intervals, comparing: 1) the NTDscope images with conventional light microscopy, both manually interpreted by microscopists, 2) the AI models with microscopist interpretation, both applied to images from the NTDscope, and 3) the AI models run on NTDscope images with conventional light microscopy. We further evaluated Pearson’s correlation coefficients for quantitative egg counts by AI models and expert microscopists on NTDscope images.

## Results

Seventy-five samples were processed on day 1 and quantified on conventional light microscopy by trained field microscopists, 25 with high-intensity infection, 25 with low-intensity infection, and 25 with no eggs present. Sixty-eight samples were processed on day 2 and again quantified on conventional light microscopy by trained field microscopists, 24 with high-intensity infection, 21 with low-intensity infection, and 23 with no eggs present.

Quantification of *S. haematobium* eggs by AI models loaded on the NTDscope was compared with the manual quantification of eggs in images acquired using the NTDscope. The results comparing the starting and edge-tuned AI models to the manual counts of eggs in NTDscope images are shown in Table 1 for brightfield and Table 2 for darkfield. Supplementary Figure 1 shows a comparison of the performance of the starting AI models on days 1 and 2 for the sensitivity-optimized and specificity-optimized thresholds. After the study was completed, we compared the starting and edge-tuned AI model performance on day 2 patient data to directly assess the model improvements that were gained by edge-tuning.

Figure 3 shows a comparison between the starting and edge-tuned AI models on day 2 for the sensitivity-optimized (Figure 3a), specificity-optimized (Figure 3b), and egg quantification- optimized thresholds (Figure 3c). For the egg quantification-optimized AI models, the changes in slope, root mean square error (RMSE), and Pearson’s r are shown for each plot. Edge-tuning resulted in noticeable improvements in performance for all thresholds and contrasts we evaluated. The edge-tuned AI models using the sensitivity-optimized threshold (Figure 3a) were the highest-performing models on day 2. For this threshold, the brightfield starting model performance increased from a sensitivity of 75.5% and specificity of 46.7% to an edge-tuned sensitivity of 98.1% and specificity of 100%. The darkfield starting model performance increased from a sensitivity of 72.2% and specificity of 64.3% to an edge-tuned sensitivity of 90.7% and specificity of 100%.

We compared the AI-based and manual counting of *S. haematobium* eggs in images acquired on the NTDscope to the egg counts done by field microscopists using conventional light microscopy. The results for manual counts of eggs in NTDscope images compared to light microscopy are shown in Table 3. The results for the AI models compared to light microscopy are shown in Tables 4-5. Figure 4 shows patient-level sensitivity and specificity of the highest performing edge-tuned AI model (using the sensitivity-optimized threshold, Figure 4a) and the manual counts of eggs in NTDscope images on day 2 (Figure 4b). On day 2, the edge-tuned brightfield AI model had a sensitivity of 97.8% and specificity of 68.2% when compared to light microscopy. The darkfield model had a sensitivity of 95.7% and specificity of 77.3%. Manual counts of eggs in NTDscope brightfield images had a sensitivity of 97.8% and specificity of 63.6% while manual counts of eggs in NTDscope darkfield images had a sensitivity of 97.8% and a specificity of 59.1%. Patient-level performance of edge-tuned AI models on the NTDscope was equivalent to human counts on the same images acquired with the NTDscope.

**Figure 4:**
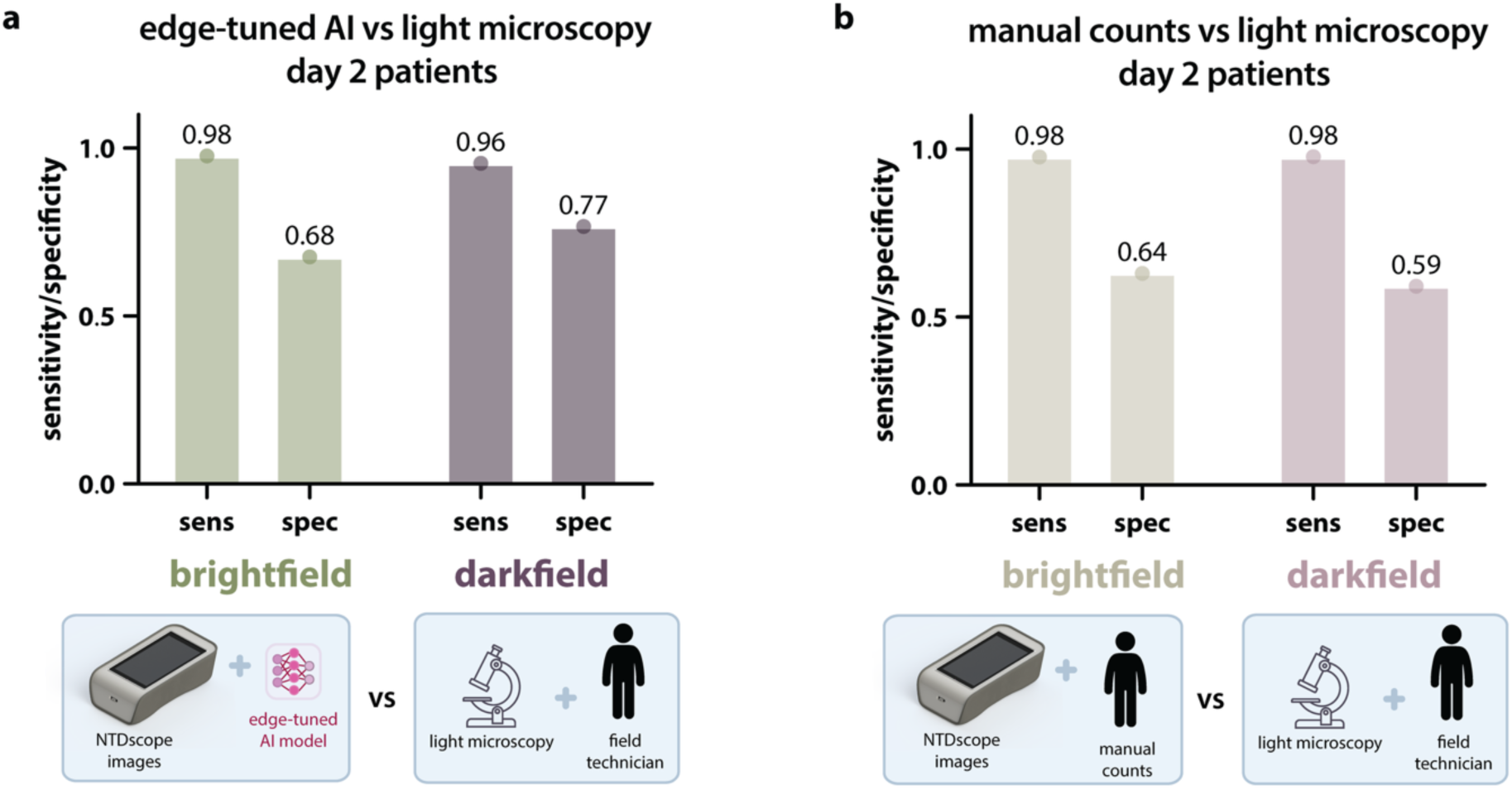
Comparison of edge-tuned AI models and manual counts of S. haematobium eggs on NTDscope images using light microscopy as the gold standard. **a.** Results from brightfield and darkfield edge-tuned AI models on the NTDscope, compared to light microscopy. These are results using the sensitivity-optimized threshold. **b.** Results from manual egg counts on NTDscope, compared to light microscopy. Edge-tuned AI models run on the NTDscope have equivalent performance to manual counts when compared to light microscopy.

**Table 3:** Performance of manual counts on NTDscope images compared to light microscopy.

| Day | Contrast | Patient-level |  |
| --- | --- | --- | --- |
|  |  | Sensitivity | Specificity |
| 1 | Brightfield | 94% | 68% |
| 1 | Darkfield | 100% | 72% |
| 2 | Brightfield | 97.8% | 63.6% |
| 2 | Darkfield | 97.8% | 59.1% |

**Table 4:** Performance of brightfield AI models compared to light microscopy.

| Day | AI model | Threshold | Patient-level |  |
| --- | --- | --- | --- | --- |
|  |  |  | Sensitivity | Specificity |
| 1 | Starting model | Egg-quantification | 96% | 56% |
| 1 | Starting model | Sensitivity-optimized | 100% | 16% |
| 1 | Starting model | Specificity-optimized | 72% | 72% |
| 2 | Starting model | Egg-quantification | 76.1% | 50% |
| 2 | Starting model | Sensitivity-optimized | 78.3% | 45.5% |
| 2 | Starting model | Specificity-optimized | 76.1% | 63.6% |
| 2 | Edge-tuned model | Egg-quantification | 97.8% | 50% |
| 2 | Edge-tuned model | Sensitivity-optimized | 97.8% | 68.2% |
| 2 | Edge-tuned model | Specificity-optimized | 84.8% | 81.8% |

**Table 5:** Performance of darkfield AI models compared to light microscopy.

|  |  |  | Patient-level |  |
| --- | --- | --- | --- | --- |
| Day | AI model | Threshold | Sensitivity | Specificity |
| 1 | Starting model | Egg-quantification | 72% | 80% |
| 1 | Starting model | Sensitivity-optimized | 92% | 40% |
| 1 | Starting model | Specificity-optimized | 40% | 88% |
| 2 | Starting model | Egg-quantification | 73.9% | 68.2% |
| 2 | Starting model | Sensitivity-optimized | 73.9% | 54.5% |
| 2 | Starting model | Specificity-optimized | 65.2% | 81.8% |
| 2 | Edge-tuned model | Egg-quantification | 95.7% | 54.5% |
| 2 | Edge-tuned model | Sensitivity-optimized | 95.7% | 77.3% |
| 2 | Edge-tuned model | Specificity-optimized | 84.8% | 86.4% |

## Discussion

We show that diagnosis of *S. haematobium* infections in the field can be improved by edge- tuning AI models running on a handheld, portable microscope, demonstrating a promising diagnostic workflow for screening in low-resource settings.

Various methods have leveraged AI to diagnose endemic infections in rural or remote regions of low- and middle-income countries (LMICs). One effective approach involves transporting samples to a central laboratory, where images are scanned and uploaded to the cloud for AI-driven analysis of specimens; this method has recently proven successful in diagnosing soil-transmitted helminths.^17^ Other solutions proposed to run AI models offline, leveraging a smartphone coupled to a traditional optical microscope in lab settings.^8^ Our approach involves performing AI analysis directly on a highly portable mobile microscope, which can reduce time and limit costs by allowing samples to be evaluated in the field at the point of sample collection.

A well-documented phenomenon in the field of AI is the dataset shift, where differences between the data used for training and testing AI models can degrade model performance.^18,19^ This impact is especially noticeable when the training dataset is small and comes from a single source. A strategy to mitigate such decreases in performance when using models in new clinical contexts is to re-train or fine-tune the models using data from the new setting. In our case, the starting AI model was trained using images from an older version of our portable microscope, with only 218 images for training and 45 for validation. Since the new NTDscope has slightly different optics and illumination^11^, and given the small dataset used for training the starting AI model, we suspected that performance could improve from re-training the model in the field with images acquired using the new NTDscope, a technique we call edge-tuning. We demonstrated that the methodology deployed in this study could be used to retrain and deploy a new version of the AI model in less than 24 hours in a low-resource setting, leveraging a pipeline including portable image acquisition, mobile connectivity, off-site cloud-based labelling and training, model evaluation and optimization, model deployment and on-the-edge inference execution. We evaluated the impact on model performance gained by edge-tuning by comparing the performance of the starting and edge-tuned AI models on the patient images on day 2, which showed robust improvements for across AI model confidence score thresholds.

When using light microscopy as a reference, our edge-tuned AI models had comparable performance to manual counts of eggs in NTDscope images for both brightfield and darkfield versions of the model (Figure 4), validating the usefulness of edge-tuned AI models for simplifying the work of field technicians performing manual egg counts. The discrepancies between conventional microscopy and the NTDscope may be due to the fact that different fractions of the same urine sample were used in each case. Running AI models on NTDscope images is considerably easier and faster than performing manual counts and does not require highly trained microscopists, who are unfortunately in short supply in many African settings; rather, image acquisition and processing on the device take <2 min per patient. To our knowledge, this is the first demonstration of re-training of an AI model for neglected tropical disease screening within one day in an endemic, low-resource setting. The combination of our devices’ portability and mobile connectivity, as well as our infrastructure for image annotation and model re-training, enabled us to quickly adapt to a new clinical setting and improve our diagnostic performance without losing significant time during the field study. Our performance compared to light microscopy is close to meeting the WHO requirements for monitoring and evaluation of schistosomiasis control programmes, and we expect that future AI models edge- tuned with additional field data will enable us to reach the required metrics.

Portable microscopy, with iterative edge-tuning under different conditions to improve AI model performance and generalizability, has significant potential to improve the quality of public health programs aimed at controlling neglected tropical infections. Such technology has already supported identifying high-intensity *Loa loa* infections, malaria, schistosomiasis, and soil- transmitted helminthiases.^5–8,13,16,17,20^ Given the scarcity of laboratory capacity and trained personnel in rural areas where schistosomiasis is endemic,^21^ an AI-powered NTDscope could expand access to essential public health programs for a greater number of individuals, especially when combined with point-of-care circulating cathodic antigen testing for *S. mansoni*; such a combined system can enable low-cost and rapid screening for both urogenital (*S. haematobium*) and gastrointestinal (*S. mansoni*) schistosomiasis by testing only urine.^13^ This ’urine-only’ approach could be a valuable method for monitoring existing schistosomiasis control and elimination programs, addressing a significant need identified by the WHO.^2^

## Data Availability

The data and algorithms may be made available upon request. Interested parties are encouraged to contact the corresponding author for further information.

## Contributors

MDDLD, ED, DC, DBP, JGV, LL, KNF, DAF, MLO, and IIB designed the research and analysis. JTC, ED, KDS, DC, KNF, and IIB collected the data. MDDLD, ED, DC, and JGV prepared the portable microscopes for data collection. MDDLD, JTC, ED, KDS, DC, DBP, JGV, LL, KNF, and IIB participated in data annotation. ED, DC, DBP, JGV, LL, and MLO trained AI models. MDDLD, DBP, LL, KNF, JRA, DAF, and IIB performed statistical analysis. All authors participated in data interpretation. MDDLD, JTC, ED, DC, DBP, JGV, LL, KNF, DAF, MLO, and IIB co-wrote the manuscript, and all authors provided feedback. All authors had full access to all the data in the study and accepted the responsibility to submit for publication.

