## Supplementary Figure 1 for "Edge-tuning of artificial intelligence improves diagnostic performance for *Schistosomiasis haematobium* in a rural setting of Côte d’Ivoire"

### Supplementary Figures

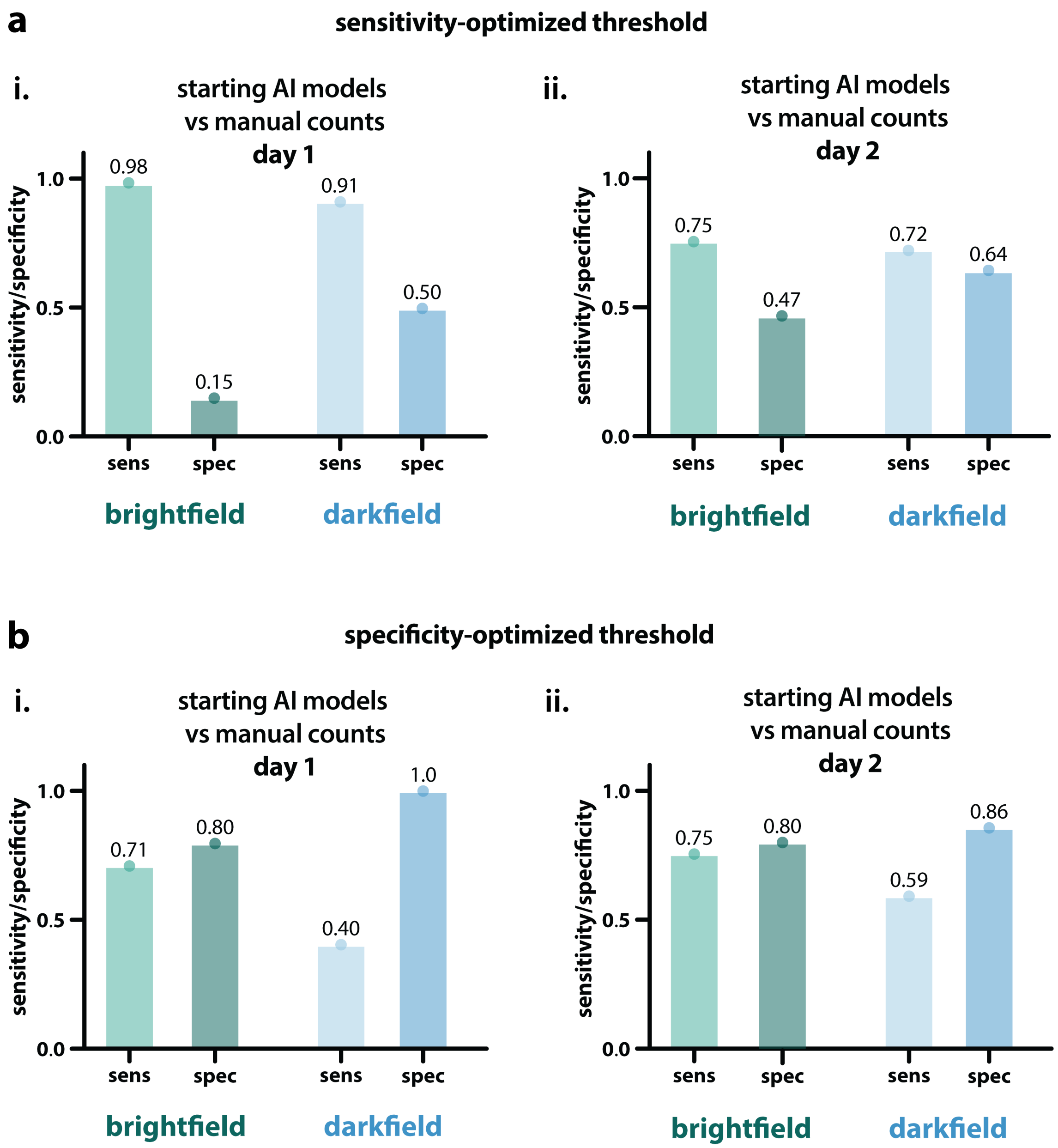


***Supplementary Figure 1**: Diagnostic performance of starting AI models compared to manual counts on NTDscope images, showing the results using sensitivity-optimized (**a**) and specificity-optimized (**b**) confidence score thresholds from field days 1 (**i**) and 2 (**ii**). These starting AI models were trained on images containing* S. haematobium *eggs collected in previous field studies in Côte d'Ivoire using a previous version of the NTDscope.*
